# Infoveillance based on Social Sensors to Analyze the impact of Covid19 in South American Population

**DOI:** 10.1101/2020.04.06.20055749

**Authors:** Josimar E. Chire Saire

## Abstract

Infoveillance is an application from Infodemiology field with the aim to monitor public health and create public policies. Social sensor is the people providing thought, ideas through electronic communication channels(i.e. Internet). The actual scenario is related to tackle the covid19 impact over the world, many countries have the infrastructure, scientists to help the growth and countries took actions to decrease the impact. South American countries have a different context about Economy, Health and Research, so Infoveillance can be a useful tool to monitor and improve the decisions and be more strategical. The motivation of this work is analyze the capital of Spanish Speakers Countries in South America using a Text Mining Approach with Twitter as data source. The preliminary results helps to understand what happens two weeks ago and opens the analysis from different perspectives i.e. Economics, Social.

## 1 Introduction

Infodemiology^1^ is a new research field, with the objective of monitoring public health^2^ and support public policies based on electronic sources, i.e. Internet. Usually this data is open, textual and with no structure and comes from blogs, social networks and websites, all this data is analysed in real time. And Infoveillance is related to applications for surveillance proposals, i.e. monitor H1N1 pandemic with data source from Twitter^3^, monitor Dengue in Brazil^4^, monitor covid19 symptoms in Bogota, Colombia^5^. Besides, Social sensors is related to observe what people is doing to monitor the environment of citizens living in one city, state or country. And the connection to Internet, the access to Social Networks is open and with low control, people can share false information(fake news)^6^.

A disease caused by a kind coronavirus, named Coronavirus Disease 2019 (covid19) started in Wuhan, China at the end of 2019 year. This virus had a fast growth of infections in China, Italu and many countries in Asia, Europe during January and February. Countries in America(Central, North, South) started with infections at the middle of February or beginning of March. This disease was declared a global concern at the end of January by World Health Organization(WHO)^7^.

South America has different context about economics, politics and social issues than the rest of the world and share a common language: Spanish. The decisions made for each government were over the time, with different dates and actions: i.e. social isolation, close limits by air, land. But, there is no tool to monitor in real time what is happening in all the country, how the people is reacting and what action is more effective and what problems are growing.

For the previous context, the motivation of this work is analyze the capitals of countries with Spanish as language official to analyze, understand and support during this big challenge that we are facing everyday.

This paper follows the next organization: section 2 explains the methodology for the experiments, section 3 presents results and analysis. Section 4 states the conclusions and section 5 introduces recommendations for studies related.

## 2 Methodology

The present analysis is inspired on Cross Industry Standard Process for Data Mining(CRISP-DM)^8^ steps, the phases are very frequent on Data Mining tasks. So, the steps for this analysis are the next:

- Select the scope of the analysis and the Social Network
- Find the relevant terms to search on Twitter
- Build the query for Twitter and collect data
- Cleaning data to eliminate words with no relevance(stopwords)
- Visualization to understand the countries

### 2.1 Selecting the scope and Social Network

Considering the countries where Spanish is the official language, there are 9 countries in South America: Argentina, Bolivia, Chile, Colombia, Ecuador, Paraguay, Perú, Uruguay, Venezuela and every nation has a different territory size as the table Tab. 1 shows.

Therefore, analyze the whole countries could take a great effort about time then the scope of this paper considers the capital1 of each country because the highest population is found there.

At the same time, there are many Social Networks with like Facebook, Linkedin, Twitter, etc. with different kind of objective: Entertainment, Job Search and so on. During the last years, data privacy is an important concern and there is update on their politics, so considering the previous restriction Twitter is chosen because of the open access through Twitter API, the API will help us to collect the data for the present study. Although, the free access has a limitation of seven days, the collecting process is performed every week.

### 2.2 Find the relevant terms to search

Actually, there is hundreds of news around the world and dozens of papers about the coronavirus so to perform the queries is necessary to select the specific terms and consider the popular names over the population. The selected terms are:

- ‘coronavirus’,’covid19’

Ideally, people only uses the previous terms but, citizens does not write following this official names then special characters are found like @, #, –, _. For this reason, variations of coronavirus and covid19 are created, i.e. {‘@coronavirus’, #covid-19’, ‘@covid_19’}

### 2.3 Build the Query and collect data

The extraction of tweets is through Twitter API, using the next parameters:

- date: 08-03-2020 to 21-03-2020, the last two weeks
- terms: the chosen words mentioned in previous subsection
- geolocalization: the longitude and latitude of every capital
- language: Spanish
- radius: 50 km

### 2.4 Preprocessing Data

- Change format of date to year-month-day
- Eliminate alphanumeric symbols
- Uppercase to lowercase
- Eliminate words with size less or equal than 3
- Add some exceptions to eliminate, i.e. ‘https’, ‘rt’

### 2.5 Visualization

This step will help to answer some question to analyze what happens in every country.

- How is the frequency of posts everyday?
- Can we trust on all the posts?
- The date of user account creation
- Tweets per day to analyze the increasing number of posts
- Cloud of words to analyze the most frequent terms involved per day

## 3 Results

The next graphics presents the results of the experiments and answer many questions to understand the phenomenon over the population.

### 3.1 What is the frequency of users posts?

At beginning, a fast preview about the frequency of post per country will support us to understand how many active users are in every capital.

Four things are important to highlight from Fig. 2: (1) Venezuela is a smaller country but the number of posts are pretty similar to Argentina, (2) Paraguay is almost a third from Peru territory and the number of publications are very similar, Chile is one small country but the number of publication are higher than Peru and (4) Uruguay is the smallest one with more tweets than Bolivia and Colombia even Ecuador has more.

**Figure 1.**
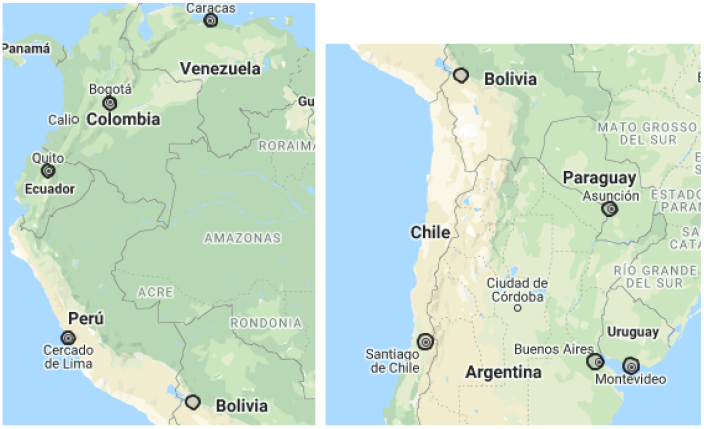
Geolocalization of South American Capital of Spanish Speakers

**Figure 2.**
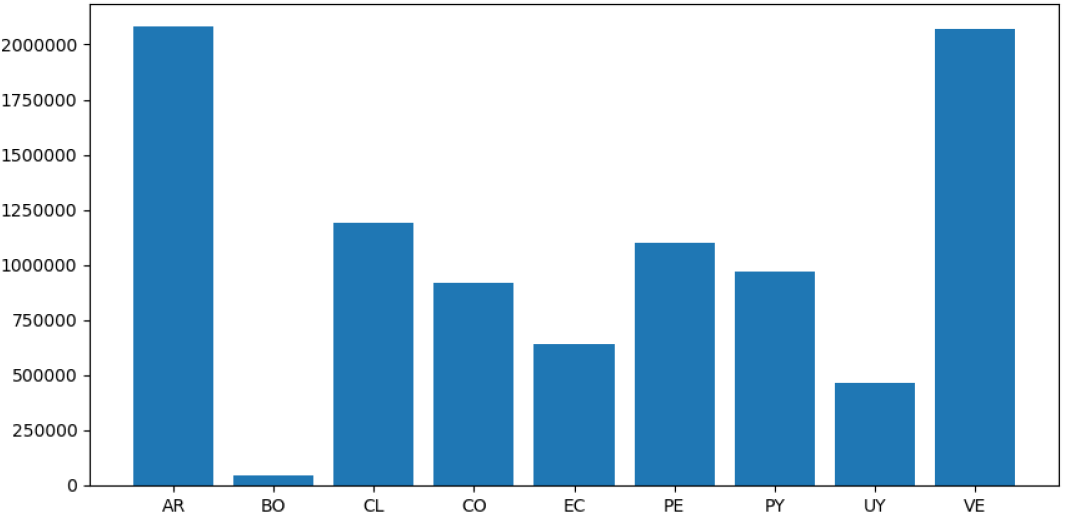
Data User Creation

By other hand, considering data from Table 1, there is a strong relationship between Internet, Social Media and Mobile Connections in Argentina, Venezuela with the number of tweets and but a different context for Colombia, this insight show us the level of using in Bogota and says how the Internet Users are spread in other cities on Colombia. So, a similar behavior explained previously is present over this data.

**Table 1.** South America Cartography Information

| Name | Capital | Territory(km) |
| --- | --- | --- |
| Argentina | Buenos Aires | 2.792.600 km |
| Bolivia | La Paz | 1.098.581 km |
| Chile | Santiago | 756.102 km |
| Colombia | Bogotá | 1.141.748 km |
| Ecuador | Quito | 283.561 km |
| Paraguay | Asunción | 406.752 km |
| Perú | Lima | 1.285.216 km |
| Uruguay | Montevideo | 176.215 km |
| Venezuela | Caracas | 916.445 km |

### 3.2 Can we trust on all this posts?

Considering the image Fig.2, the number of post for each country, the total number of tweets is up to five millions(5 627 710), close to half of million(401 979) per day. So, the question about veracity is important to filter and analyze what people is thinking, because the noise could be a limitation to understand what truly happens. By consequence, it is necessary to consider some criterion to filter this data.

#### 3.2.1 Who are the 100 top users in every country?

First, Argentina has the highest number of publications in last two weeks. For example, the firt dozen of the top users in Buenos Aires are:

‘Portal Diario’, ‘.’, ‘Clarín’, ‘Radio DoGo’, ‘Camila’,’El Intransigente’, ‘Agustina’, ‘Pablo’, ‘FrenteDeTodos’, ‘Ale’,’Lucas’, ‘Diario Crónica’ Later, a search about the users, one natural finding is: they are related to newspapers, radio or television(mass media). But there is people with many hundreds of tweets and regular people. The next image Fig. 3 has the names of users and quantity of posts.

**Figure 3.**
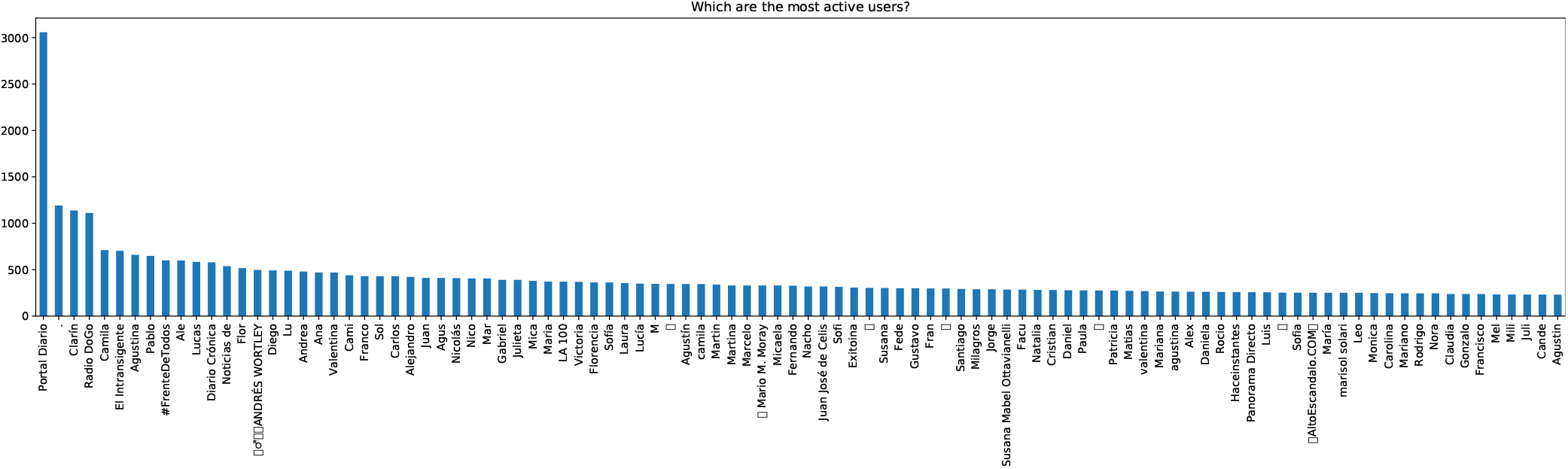
First 100 users with more posts

This users has an average of half thousand of tweets in two weeks, around 35 tweets per day each one. Selecting this people, the number of publication decreases drastically, see Fig.4

**Figure 4.**
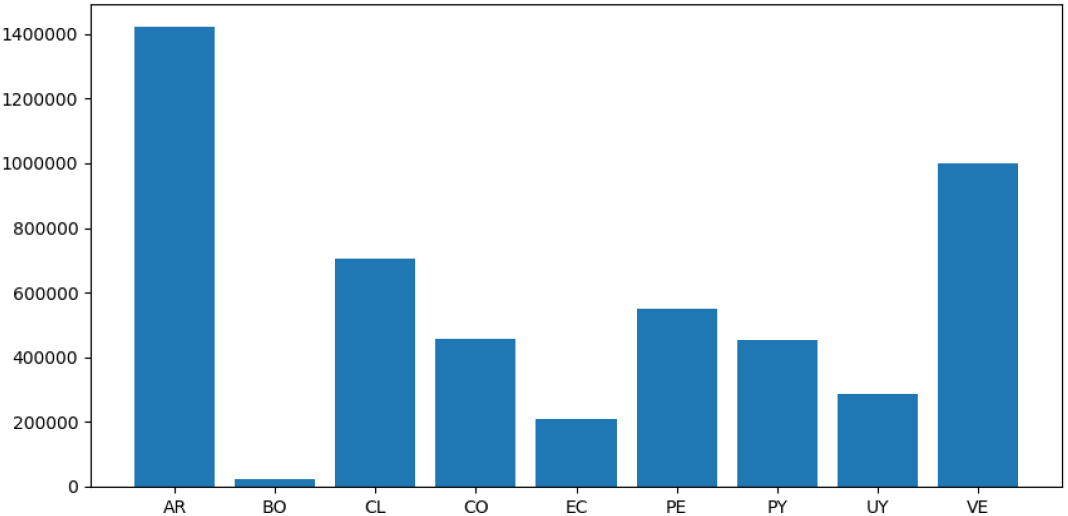
Number of Tweets per Country

A similar scenario is present in all the countries, mass media is part of the top users and regular people is posting, so it is possible to know what they are thinking about covid19. Table Tab. 3 introduces a summary of the top users per country.

**Table 2.** Access Internet Information^9^(millions)

| Name | Internet Users | Social Media Users | Mobile Connections |
| --- | --- | --- | --- |
| Argentina | 35.09 | 34.00 | 58.21 |
| Bolivia | 7.50 | 7.50 | 11.48 |
| Chile | 15.67 | 15.00 | 26.32 |
| Colombia | 35.00 | 35.00 | 60.38 |
| Ecuador | 12.00 | 12.00 | 15.65 |
| Paraguay | 4.61 | 4.00 | 7.21 |
| Perú | 24.00 | 24.00 | 38.08 |
| Uruguay | 2.70 | 2.70 | 5.42 |
| Venezuela | 20.50 | 12.00 | 23.21 |

**Table 3.**
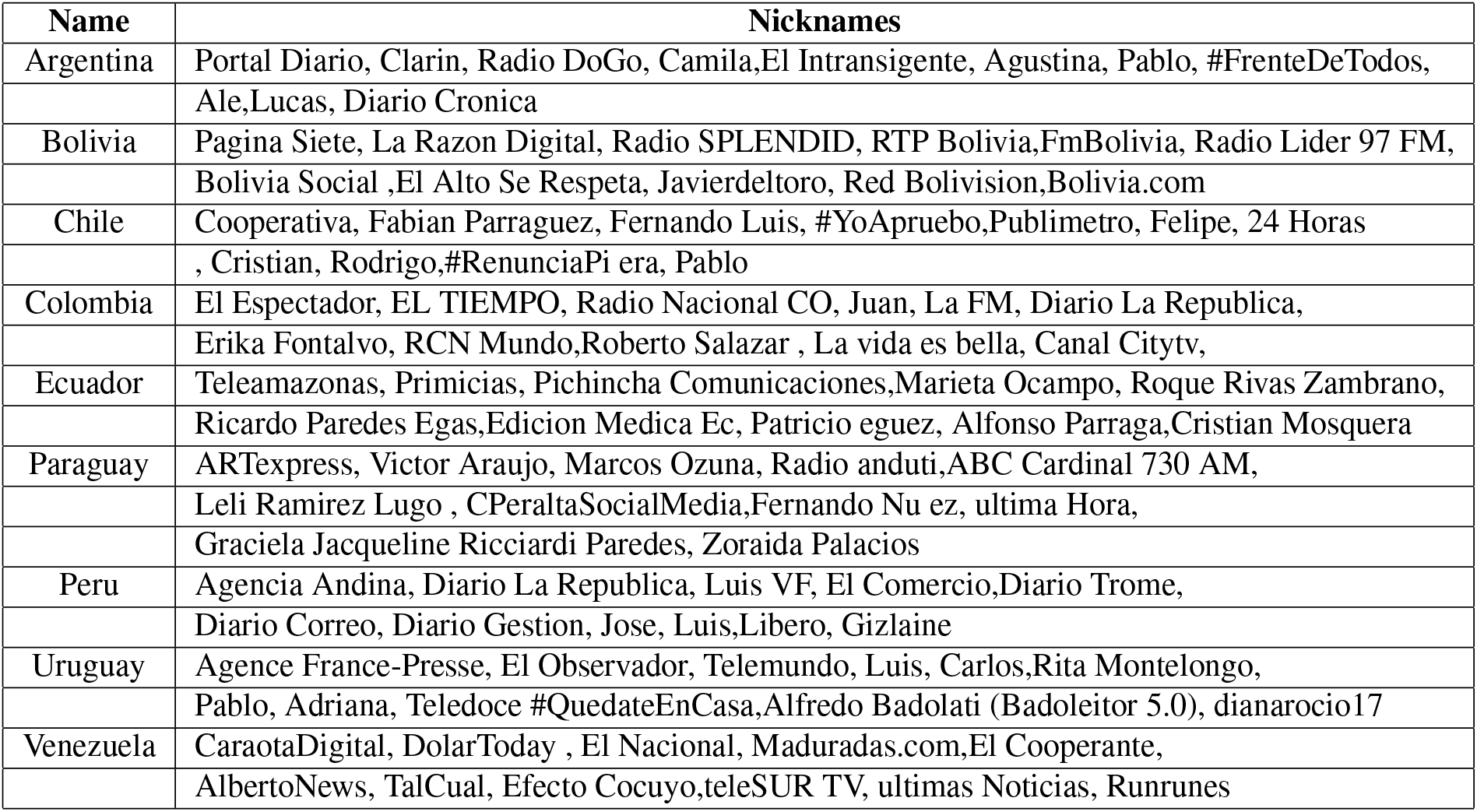
Eleven nicknames from the top 100 users

Usernames do not follow grammar rules so alphanumeric characters are found even characters from other languages as: Hebrew, Korean, Russian a three names with this alphabet.

### 3.3 What is the people posting about Covid19 on Twitter?

Analyzing La Paz, Bolivia Fig.5, the one hundred of more frequent terms are related to cases of coronavirus in Bolivia and extracting a value between number of tweets and number of time for each team is natural to conclude the most important topic is related to health and covid19.

**Figure 5.**
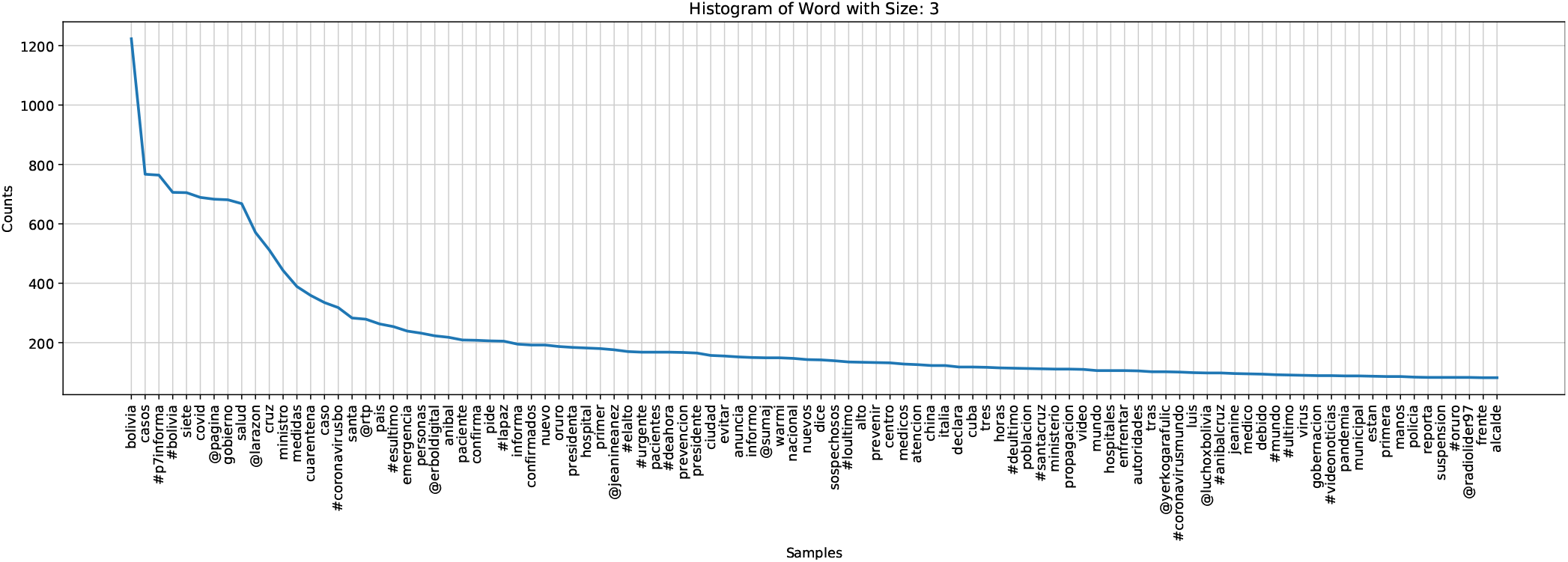
Word Histogram with size 3

Helping the visualisation from Monday to Sunday during the last two weeks, a cloud of words is presented in Fig. 6 showing the first thirty terms per country. It is important to remember every country promote different actions on different dates.

**Figure 6.**
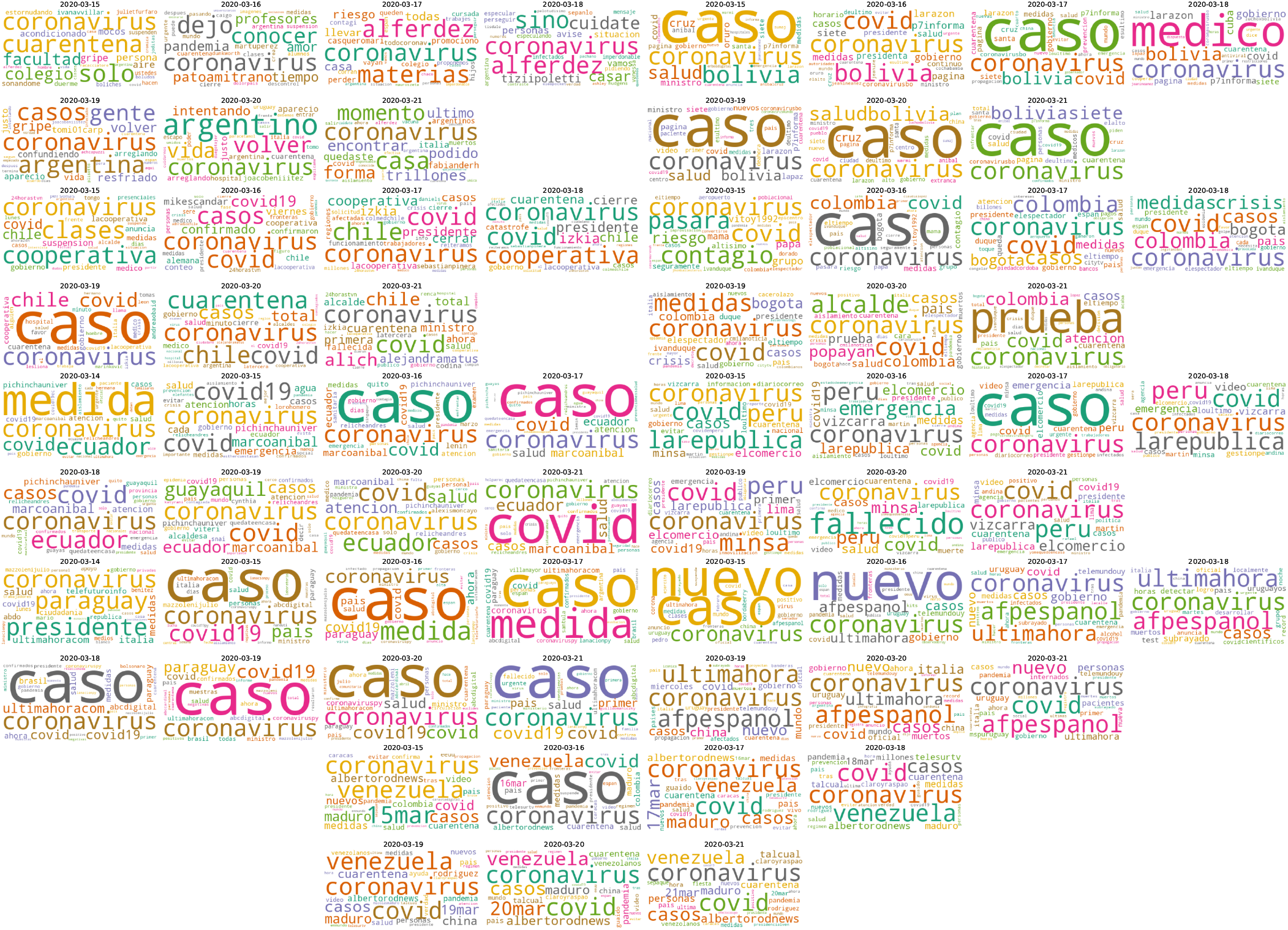
Cloud of Words: Argentina, Bolivia, Chile, Ecuador, Peru, Paraguay, Uruguay, Venezuela(left to right)

## 4 Conclusions

Infoveillance based on Social Sensors with data coming from Twitter can help to understand the trends on the population of the capitals. Besides, it is necessary to filter the posts for processing the text and get insights about frequency, top users, most important terms. This data is useful to analyse the population from different approaches.

## Data Availability

The data is available through email contact and It will be shared on github.com/jecs89 repo.

## Notes

### Competing Interest Statement

The authors have declared no competing interest.

### Funding Statement

No external funding.

